# Circulating Inflammatory Cytokines and Risk of Intracranial Aneurysm: A Mendelian-randomization Study

**DOI:** 10.1101/2023.12.17.23300093

**Authors:** Quan Zhang, Tiantian Liu, Linlin Zhang, Rongcheng Yang, Yao Zhao, Xiaojing Sun, Ting Wang, Xiaoxiao Kuang, Hengfang Liu, Jiao Liang, Ya Liu, Tao Feng

## Abstract

**Background:** Multiple studies have established the significant influence of inflammatory factors on the initiation and advancement of intracranial aneurysm (IA). However, the causality between specific inflammatory cytokines and IA remains uncertain. This study aims to elucidate it using Mendelian randomization (MR).

**Methods:** The genetic data of cytokines were collected from a genome-wide association study (GWAS) conducted on circulating cytokines in the Finnish population, involving the genetic characteristics of 41 cytokines obtained through a meta-analysis of 8,293 samples. The genetic data for IA were derived from the most extensive GWAS study, which included 7,495 samples from individuals of European ancestry and 71,934 healthy control subjects. The inverse variance weighted method primarily determined the causal correlation between exposures and outcomes. In addition, other robust MR analysis methods and sensitivity analyses were utilized to guarantee the accuracy of the results.

**Results:** Our results indicate a negative correlation between IP-10 and unruptured intracranial aneurysms (UIA) (odds ratio, OR: 0.63, 95%CI: 0.45-0.9, p=0.01). IL-10 and VEGF exhibit a positive correlation with aneurysmal subarachnoid hemorrhage (aSAH) (OR: 1.19, 95%CI: 1.02-1.39, p=0.024; OR: 1.12, 95%CI: 1.01-1.23, p=0.025). The levels of FGFBasic and TRAIL exhibit a negative correlation with aSAH. (OR: 0.62, 95%CI: 0.42-0.92, p=0.018; OR: 0.89, 95%CI: 0.8-1, p=0.041). The increased levels of IL-12p70, IL-4, IFNg, IL-10, IL-17, IL-1ra, IL-6, IL-9, VEGF, and FGFBasic were observed as the consequence of IA. (Beta: 0.18, 95%CI: 0.08-0.28, p=6.2e-4; Beta: 0.15, 95%CI: 0.07-0.23, p=2e-4; Beta: 0.15, 95%CI: 0.07-0.23, p=3.5e-4; Beta: 0.13, 95%CI: 0.05-0.21, p=1.4e-3; Beta: 0.1, 95%CI: 0.02-0.18, p=0.015; Beta: 0.17, 95%CI: 0.02-0.32, p=0.031; Beta: 0.14, 95%CI: 0.05-0.32, p=0.003; Beta: 0.13, 95%CI: 0.01-0.24, p=0.035; Beta: 0.14, 95%CI: 0.05-0.23, p=0.002; Beta: 0.09, 95%CI: 0.01-0.17, p=0.032).

**Conclusion:** Our study suggests that IP-10 may serve as a protective factor for UIA. FGFBasic and TRAIL have demonstrated a defensive impact against aSAH, while IL-10 and VEGF have been identified as factors that elevate the risk of IA rupture. Additionally, the levels of IL-12p70, IL-4, IFNg, and various cytokines are increased due to IA pathogenesis, and these cytokines may have the function of controlling inflammation in the progression of IA.

## 1. Introduction

Saccular unruptured intracranial aneurysms (UIAs) are mainly distributed in 3%-5% of the adult population, characterized by the expansion of the affected arterial wall in the brain (1). The rupture of intracranial aneurysm (IA) can result in aneurysmal subarachnoid hemorrhage (aSAH), a severe and fatal form of hemorrhagic stroke. More than half of the patients experience a loss of self-care ability, and one-third of them do not survive (2). The commonly established factors contributing to the onset of IA mainly consist of hypertension, smoking, excessive alcohol consumption, atherosclerosis, and hemodynamic alterations. Furthermore, it has been discovered that inflammatory cytokines play a vital role in controlling the formation and advancement of IA (3). Cerebral aneurysm formation occurs as a result of mechanical stretching and hemodynamic stimulation, which leads to damage of the endothelial cells in the intima layer. This damage causes the influx of pro-inflammatory macrophages and lymphocytes into the vascular wall. In addition, the phenotype of vascular smooth muscle cells (VSMCs) in the media layer undergoes a transformation into the pro-inflammatory phenotype (4). These modifications result in the hastened advancement of inflammation in the affected arterial wall, which prompts the remodeling of the aneurysm wall and triggers the initiation and rupture of IA (5).

Most studies have demonstrated a cascade amplification effect mediated by inflammatory cells and cytokines in the inflammatory process of IA (6). The regulation of cytokines is crucial for maintaining the delicate equilibrium between pro-inflammatory and anti-inflammatory states in the human immune system (7). Hence, the utilization of cytokines is of great significance in treating IA. Nevertheless, the causal correlation between specific cytokines and IA remains uncertain. Previous research has sought to clarify the association between specific cytokines and IA. However, these observational investigations may be susceptible to unanticipated confounders and reverse causal relationships, making it challenging to establish an accurate causal correlation (8).

Mendelian randomization (MR) is a reliable and rigorous statistical method that employs single nucleotide polymorphisms (SNPs) associated with particular traits as instrumental variables (IVs). MR analysis permits meticulous management of potential confounding factors, facilitating the examination of the correlation between gene-predicted exposures and outcomes independent of environmental or lifestyle influences. Due to the random distribution of alleles during meiosis, MR analysis filters out common confounders, subjective bias, and reverse causality, providing an accurate causality assessment (9). Our study extracted genetic instruments for 41 cytokines and different types of IA from previously published genome-wide association studies (GWASs). This approach enabled us to investigate the causality between cytokines and the risk of UIA or aSAH. Additionally, we investigated the correlation between the IA and inflammatory cytokines using reverse MR analysis.

## 2. Materials and Methods

### 2.1 Study design

Three crucial assumptions underpin the efficacy of MR analysis: 1) the genetic variations identified as IVs are significantly related to assumed exposures; 2) IVs are independent of confounders in the exposure-outcome association; 3) IVs are not directly related to the outcome but solely influence the outcome through the associated exposure (10). The genetic instruments utilized in this study were derived from previously published GWAS summary data. Written informed consent from all participants was obtained and approved by the ethics committee of each institutional review board in the original studies, eliminating the need for additional ethical approval and informed consent. Our study aims to investigate the correlation between inflammatory cytokines and various stages of IA, as well as the disruption of cytokine balance resulting from the pathogenesis of IA (Figure 1).

**Figure 1.**
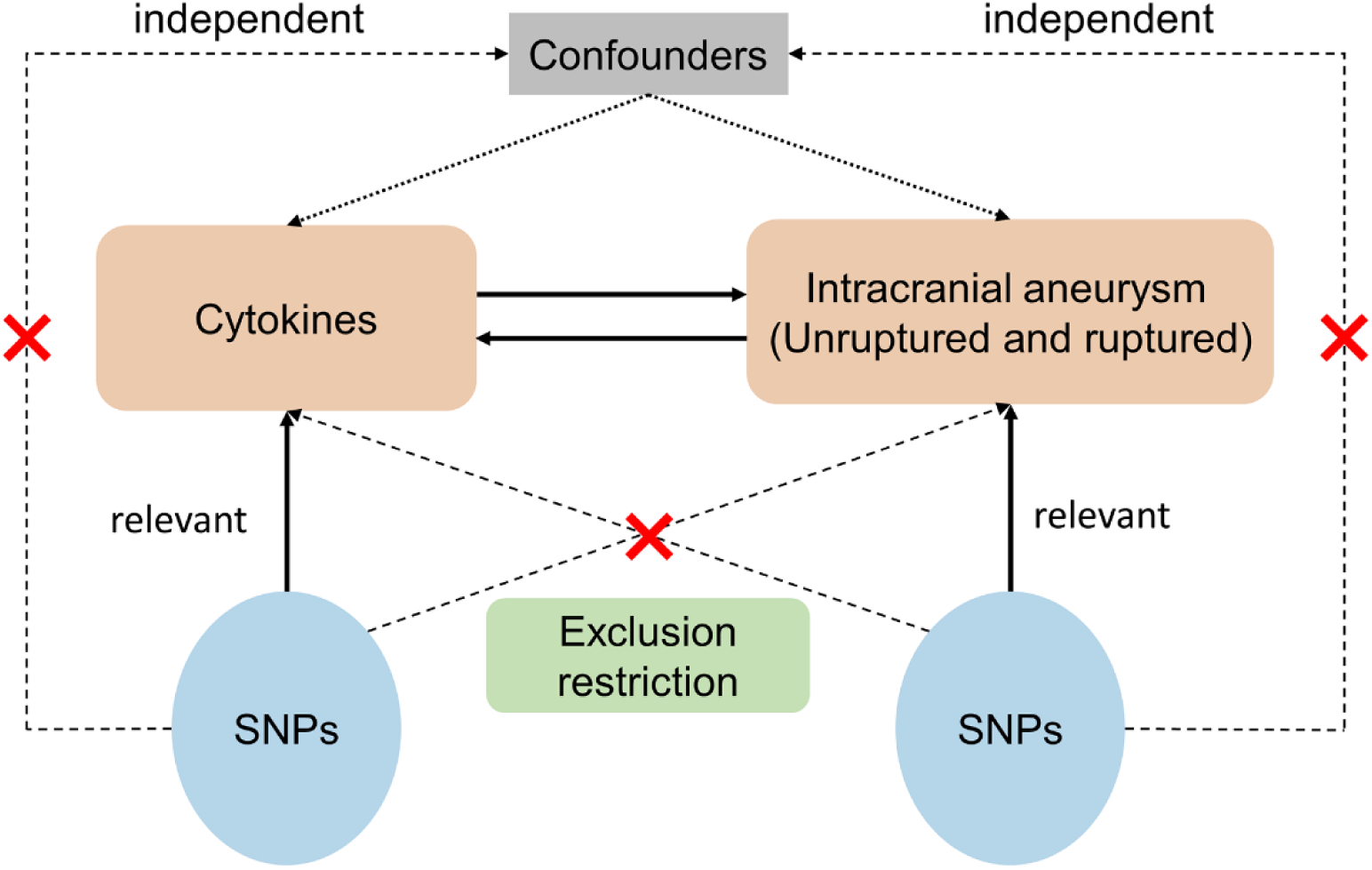
Schematic representing the study design. SNPs were selected from cytokines and intracranial aneurysms as Instrumental variables. The directed acyclic plot visually demonstrates the independence, relevance, and exclusion restriction.

### 2.2 Data source

The summary data on inflammatory cytokines were derived from a GWAS meta-analysis that included the Cardiovascular Risk in Young Finns Study (YFS) and the FINRISK studies (FINRISK 2002 and FINRISK 1997). This GWAS analyzed the cytokines genome variant associations in 8,293 Finnish adults, fitting age, gender, and body mass as covariates in the model (11). The IA summary data were obtained from a GWAS meta-analysis conducted by the International Stroke Genetics Consortium (ISGC). This study included 7,495 cases of IA (of any type), 2,070 cases of UIA, 5,140 cases of aSAH, and 71,934 healthy individuals of European descent serving as controls. These data provide a comprehensive overview of the genome variant associations of IA at each stage (12). Notably, the populations contributing cytokines and IA data were distinct.

### 2.3 Instrumental variable selection

At first, we screened cytokines and IA summary data by applying a threshold of p<5×10^-8^ to find SNPs with significant genome-wide associations. However, the initial screening did not obtain sufficient cytokine SNPs, and none of the cytokines had more than three IVs to support the study, save for MCP1. Therefore, we applied a more lenient screening threshold of p<5×10^-6^. Subsequently, we assessed the independence of all SNPs with a threshold of r^2^ < 0.001 and a clumped window of 10,000 kb to mitigate linkage disequilibrium among IVs. Given the absence of effect allele frequency information in cytokines summary data, determining the consistency of mutation direction for palindromic SNPs in the exposure and outcome was unfeasible. Thus, we excluded all the palindromic SNPs. We also calculated the F-statistic for each IV to evaluate the statistical power, employing criteria of F < 10 to identify and exclude weak IVs (13,14). Adhering to the three fundamental assumptions of MR, we utilized Phenoscanner V2 (http://www.phenoscanner.medschl.cam.ac.uk/) to reduce confounding effects. The parameters were adjusted at p = 1e-5 and r^2^ = 0.8 to exclude the SNPs associated with confounders (hypertension, smoking, and heavy drinking) (3). Finally, we excluded exposures with fewer than three SNPs from the study to support the analysis and sensitivity testing, unless sufficient proxy SNPs (R^2^ > 0.9) were added using LDlink (https://ldlink.nci.nih.gov/).

### 2.4 Statistical analysis

In this study, we employed five MR analysis methods to ensure the credibility of estimates. Under the assumption that all SNPs serve as valid instrumental variables, the inverse variance weighted (IVW) method is considered the primary criterion for establishing causality due to its highest statistical efficiency. However, this assumption cannot be definitively ascertained in the practical situation (8). Therefore, we incorporated other robust MR analysis methods into the study to provide more references for causality evaluation. The Weighted median method provides reliable MR estimates when the invalid IVs occupy less than half the weight (15). Provided the Instrument Strength Independent of Direct Effect (InSIDE) hypothesis, the MR-Egger analysis delivers consistent estimates of causal effect, mitigating the limitation of the independence assumption. Furthermore, the Simple and Weighted Mode analyses serve as supplementary references for MR results.

Considering potential horizontal pleiotropy and heterogeneity, we performed sensitivity analyses on the genetic instruments. The significance of the MR-Egger regression intercept was utilized as a criterion to assess horizontal pleiotropy (with p < 0.05) (16). In addition, we employed the MR-PRESSO approach to detect horizontal pleiotropy (with global test p < 0.05). Once the outliers identified by MR-PRESSO have been excluded, the analytic strategy will be reconsidered (17). We used the Cochran Q statistic from IVW to determine the heterogeneity of SNPs (with p < 0.05) and calculated the I^2^ statistic for a more intuitive representation. The IVW (multiplicative random effects) served as the primary method for MR analysis when significant heterogeneity among SNPs was observed without evidence of pleiotropy. The F-statistics of cytokine IVs were estimated from summary-level data. We considered weak IV bias unlikely to exist when F > 10.

Finally, we implemented the Bonferroni correction based on the number of inflammatory cytokines included in the MR analysis (p < 0.05 / N, N: the number of groups examined). The results of suggestive associations were considered significant before correction (p < 0.05) but did not retain significance after Bonferroni correction (p > 0.05 / N) (18). To assess the stability of the MR analysis, we conducted a leave-one-out sensitivity analysis to evaluate the impact of excluding any single SNP. The TwoSampleMR (19), MR-PRESSO (17), and the ggplot2 package in R (version 4.1.3) were utilized for data analysis and visualization in this study.

## 3. Results

### 3.1 The effect of inflammatory cytokines on UIA

Thirty-three inflammatory cytokines had sufficient IVs after being screened, with the cutoff value of p < 5e-6. Subsequently, we used proxy SNPs for SDF1a and TNFa to supplement the gaps caused by missing SNPs in the outcome summary data. Four SNPs (rs385076, rs57786342, rs1333040, rs635634) associated with confounders were removed by examination of phenoscanner V2. Ultimately, 35 inflammatory cytokines were included in the MR analysis. The F-statistic of each IV exceeds 10, suggesting the absence of weak IVs. After conducting the sensitivity analysis, we found no horizontal pleiotropic or heterogeneity among SNPs (Supplementary Table S1, S2).

Figure 2 depicts the association between circulating inflammatory cytokines and UIA. We observed a suggestive negative correlation between circulating levels of IP-10 and the risk of UIA (OR: 0.63, 95% CI: 0.45-0.9, p = 0.01). The weighted median method also provided a consistent assessment (OR: 0.65, 95% CI: 0.43-0.98, p = 0.039). Although the MR-Egger method did not yield significant results, it showed a similar estimate trend (OR: 0.85, 95% CI: 0.48-1.49, p = 0.67) (Supplementary Table S3). Figure 3 presents the scatter plot and funnel plot derived from MR analysis of IP-10 in UIA.

**Figure 2.**
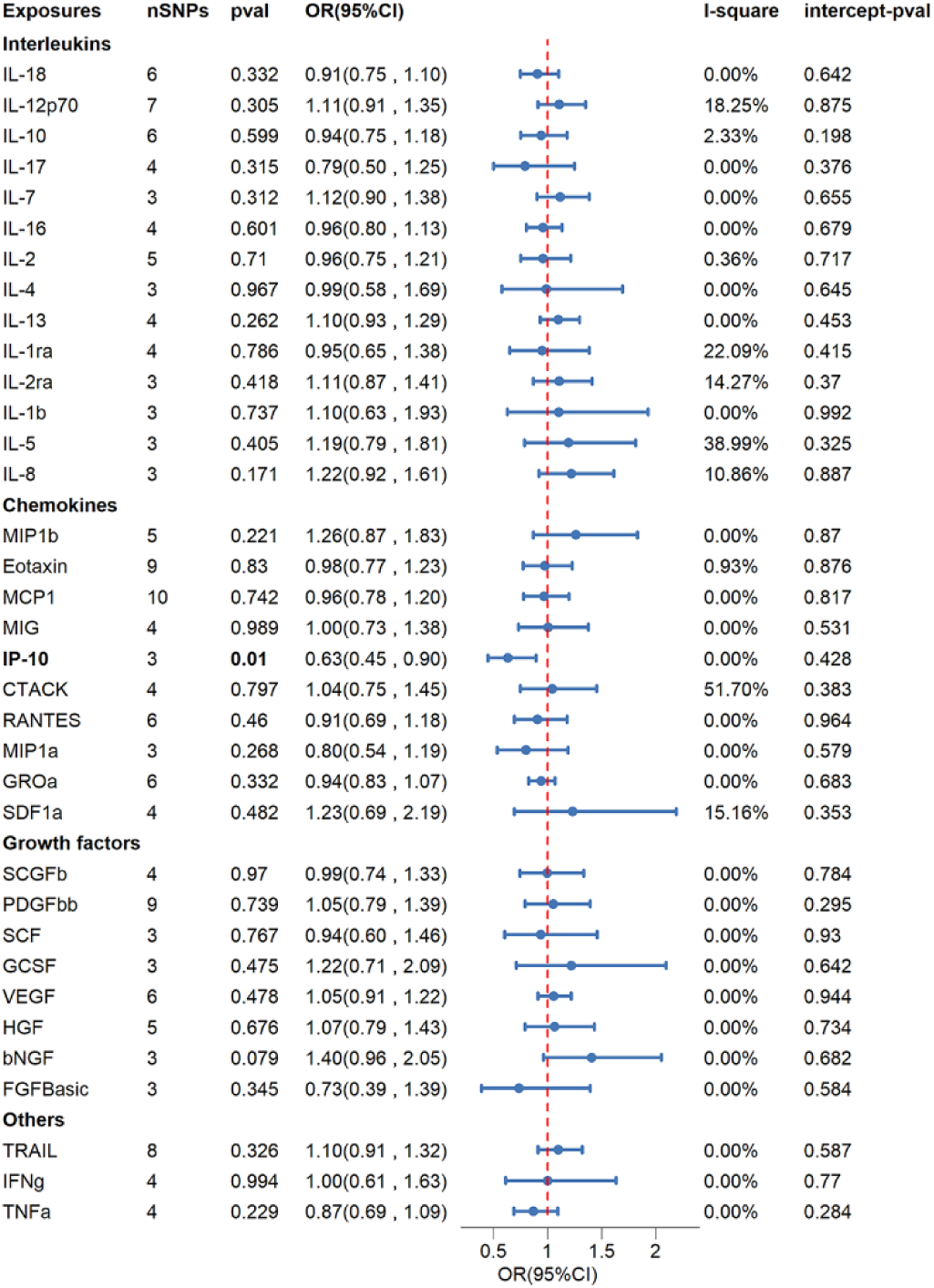
Causalities of 35 inflammatory cytokines on unruptured intracranial aneurysm (UIA). The odds ratio (OR) and 95 % confidence interval (CI) represent the odds ratio of UIA changes with per 1-SD increase in cytokines. Significance was determined using a p-value threshold of 0.05 / 35 = 0.0014 following Bonferroni correction. The inverse variance weighted method was applied consistently across all groups.

**Figure 3.**
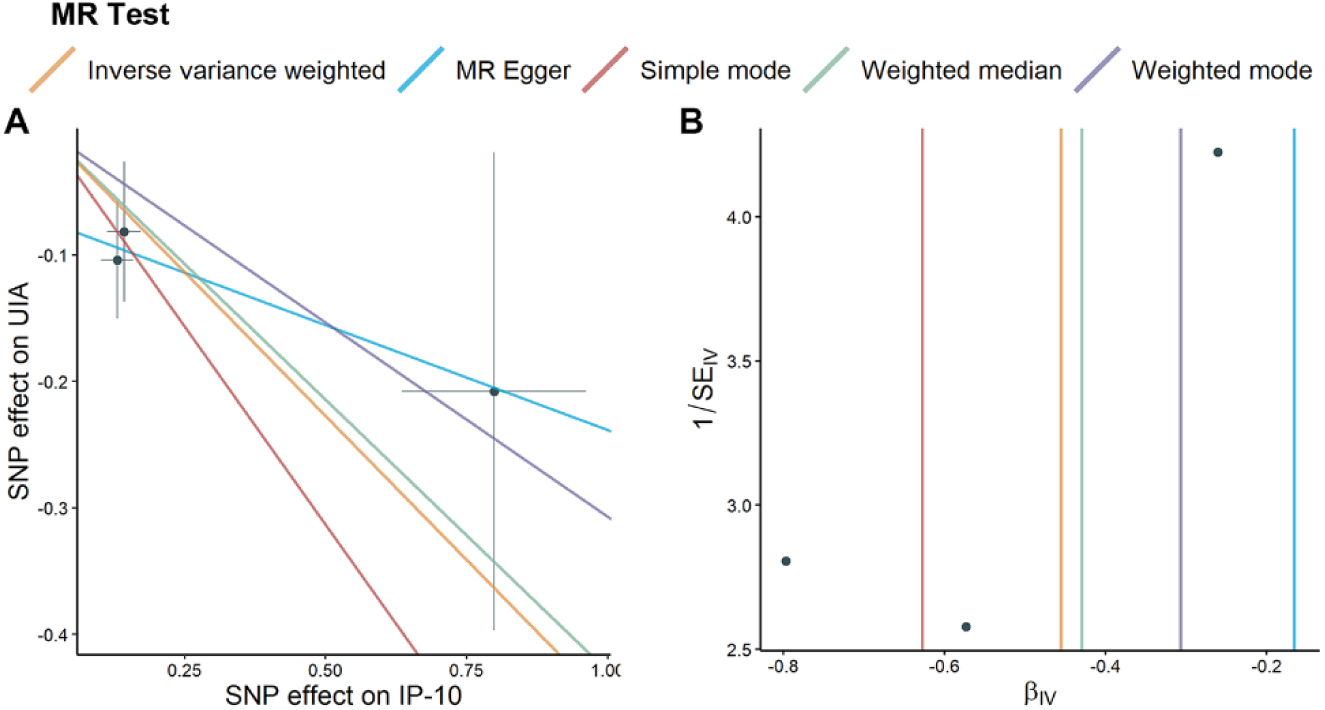
The Mendelian randomization (MR) analysis for IP-10 in unruptured intracranial aneurysms (UIA) is depicted through scatter and funnel plots. (A) Each single nucleotide polymorphism (SNP) is depicted as a black dot, with standard error bars representing its estimates of IP-10 level and UIA risk. The causality is represented by the slope of the line. (B) The MR estimation of IP-10 SNPs with UIA versus the reciprocal of the standard error (1/SEIV).

### 3.2 The effect of inflammatory cytokines on aSAH

Similarly, 35 inflammatory cytokines were included in the MR analysis after pretreatment. Among them, SDF1a and TNFa utilized proxy SNPs. Four SNPs were excluded due to their association with confounding factors (rs7088799, rs385076, rs1333040, rs635634), and no weak IV was identified. The sensitivity analysis denied the presence of horizontal pleiotropy in SNPs. Except for the IL13-aSAH group, which applied the IVW (multiplicative random effects) method due to observed heterogeneity, the other groups exhibited no heterogeneity (Supplementary Table S4, S5).

Figure 4 depicts the correlation between genetically predicted cytokines and aSAH. We observed that levels of IL-10 and VEGF were suggestively positively correlated with the risk of aSAH (OR: 1.19, 95% CI: 1.02-1.39, p = 0.024; OR: 1.12, 95% CI: 1.01-1.23, p = 0.025). Interestingly, after further MR analysis with aSAH as the exposure, we discovered the bilateral causalities between IL-10, VEGF, and aSAH (Supplementary Figure S1). In contrast, the circulating levels of FGFBasic and TRAIL were suggestively negatively associated with the risk of aSAH (OR: 0.62, 95% CI: 0.42-0.92, p = 0.018; OR: 0.89, 95% CI: 0.8-1, p = 0.041).

**Figure 4.**
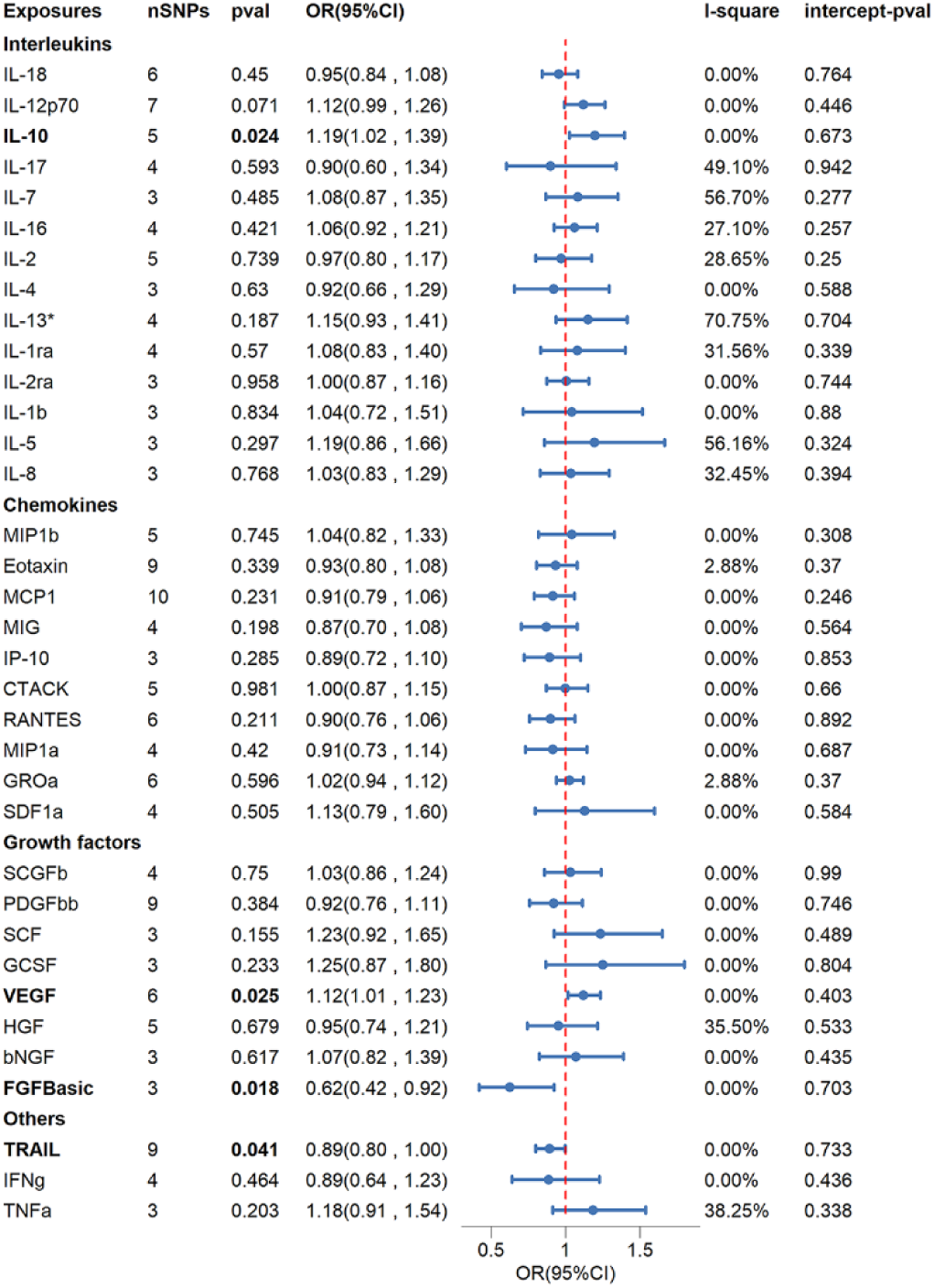
Causalities of 35 inflammatory cytokines on aneurysmal subarachnoid hemorrhage (aSAH). The odds ratio (OR) and 95 % confidence interval (CI) represent the odds ratio of aSAH changes per 1-SD rise in cytokines. Significance was determined using a p-value threshold of 0.05 / 35 = 0.0014 following Bonferroni correction. The inverse variance weighted method was applied consistently across all groups except for the IL13-aSAH group, which applied the IVW (multiplicative random effects) method.

The correlation between VEGF and aSAH was suggestively significant in the Weighted Median method (OR: 1.14, 95% CI: 1.02-1.3, p = 0.016) but not in the MR-Egger method (OR: 1.19, 95% CI: 1.01-1.4, p = 0.106). Correlations between IL-10, FGFBasic, TRAIL, and aSAH were not significant in the MR-Egger method (OR: 1.12, 95% CI: 0.83-1.52, p = 0.515; OR: 0.41, 95% CI: 0.08-2.17, p = 0.485; OR: 0.88, 95% CI: 0.77-1.01, p = 0.106) and the Weighted Median method (OR: 1.17, 95% CI: 0.99-1.39, p = 0.07; OR: 0.65, 95% CI: 0.4-1.04, p = 0.073; OR: 0.89, 95% CI: 0.78-1.02, p = 0.084). However, both methods exhibited a similar estimate trend to the IVW method (Supplementary Table S6). The scatter plots and funnel plots of the MR analysis of IL-10, VEGF, FGFBasic, and TRAIL in aSAH are presented in Figure 5 and Figure 6.

**Figure 5.**
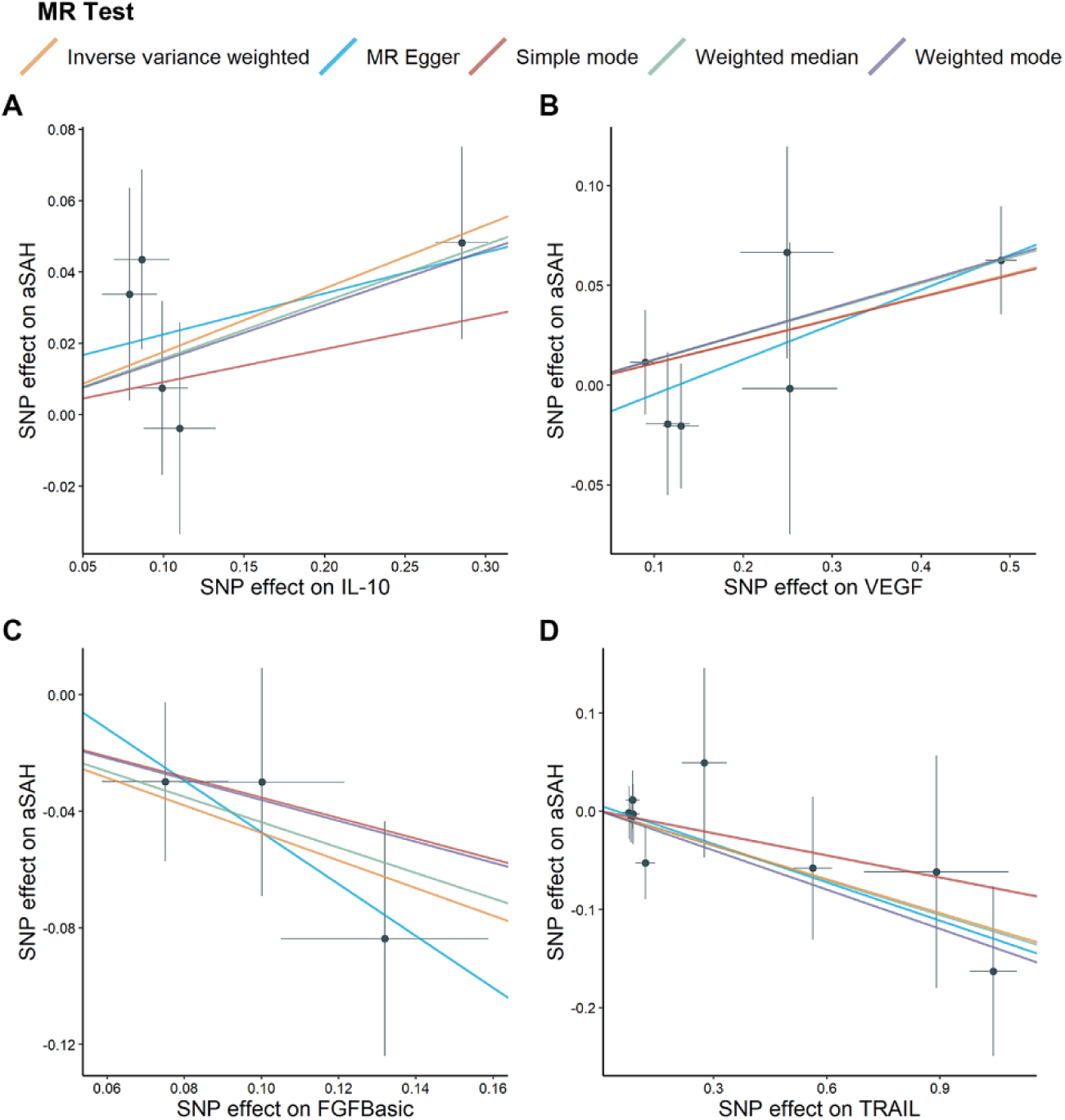
The Mendelian randomization (MR) analyses for specific cytokines in aneurysmal subarachnoid hemorrhage (aSAH) are depicted through scatter plots. Each single nucleotide polymorphism (SNP) is depicted as a black dot, with standard error bars representing its estimates of cytokine level and aSAH risk. The causality is represented by the slope of the line. (A-D): IL-10, VEGF, FGFBasic, and TRAIL.

**Figure 6.**
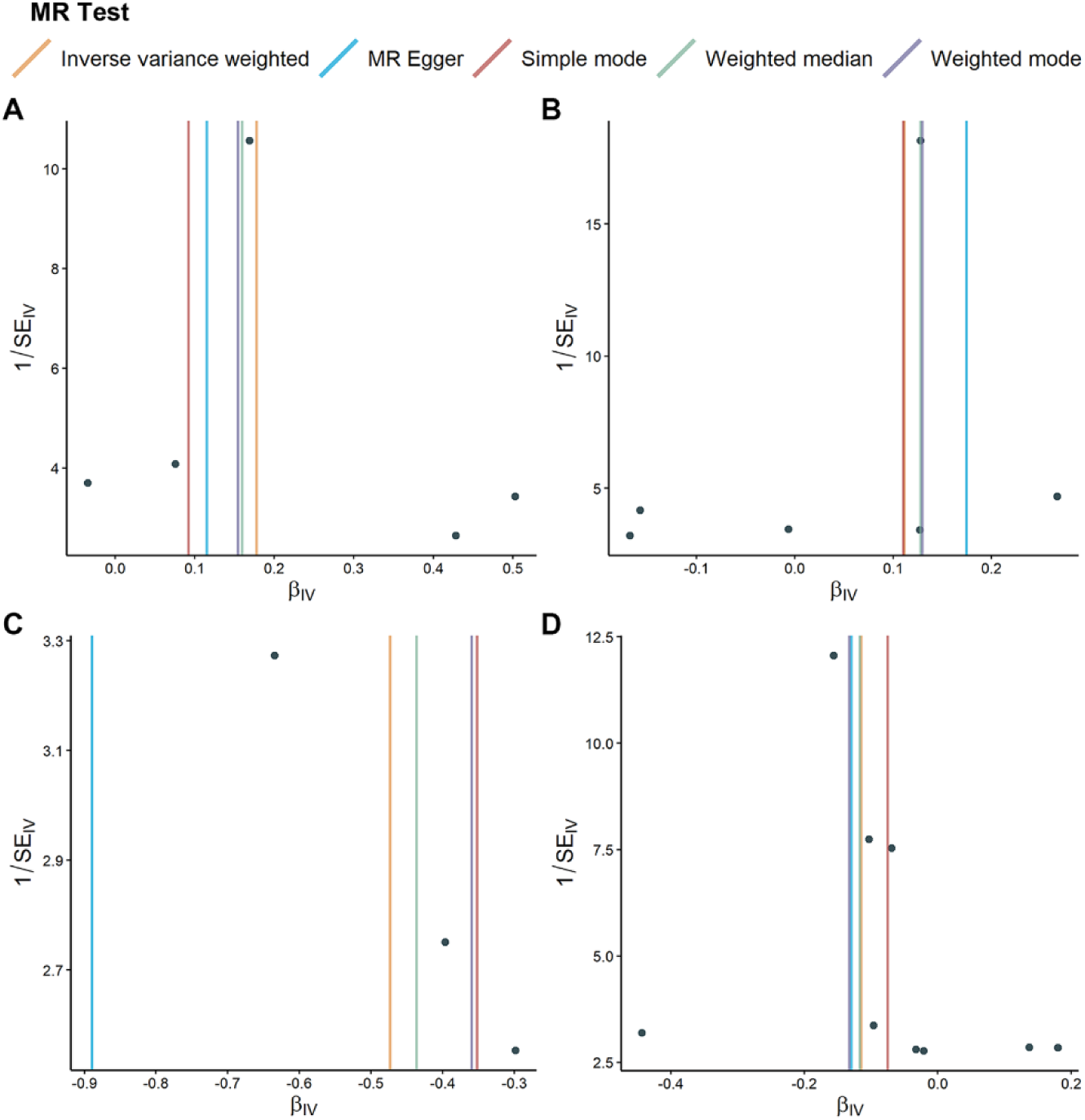
The Mendelian randomization (MR) analyses for specific cytokines in aneurysmal subarachnoid hemorrhage (aSAH) are depicted through funnel plots. The MR estimation of cytokines single nucleotide polymorphisms with aSAH versus the reciprocal of the standard error (1/SEIV). (A-D): IL-10, VEGF, FGFBasic, and TRAIL.

### 3.3 The effect of IA on cytokines

After screening SNPs in the summary data of IA with a threshold of p < 5e-8, we excluded four genetic instruments related to confounders (rs10519203, rs72841270, rs4705938, rs11646044). Ultimately, seven IVs were included in the MR analysis. F-statistics of each IV indicated a robust assumed association (median, 59.51; range, 30.15-94.24). Sensitivity analysis refutes the existence of heterogeneity and pleiotropy (Supplementary Table S7, S8).

Figure 7 illustrates the causality between IA and inflammatory cytokines. Our results indicate significant correlations between the genetically predicted IA and elevated circulating levels of IL-12p70, IL-4, and IFNg (Beta: 0.18, 95% CI: 0.08-0.28, p = 6.2e-4; Beta: 0.15, 95% CI: 0.07-0.23, p = 2e-4; Beta: 0.15, 95% CI: 0.07-0.23, p = 3.5e-4). Additionally, IA has suggestive significant positive correlations to seven other inflammatory cytokines, namely IL-10 (Beta: 0.13, 95% CI: 0.05-0.21, p = 1.4e-3), IL-17 (Beta: 0.1, 95% CI: 0.02-0.18, p = 0.015), IL-1ra (Beta: 0.17, 95% CI: 0.02-0.32, p = 0.031), IL-6 (Beta: 0.14, 95% CI: 0.05-0.32, p = 0.003), IL-9 (Beta: 0.13, 95% CI: 0.01-0.24, p = 0.035), VEGF (Beta: 0.14, 95% CI: 0.05-0.23, p = 0.002), and FGFBasic (Beta: 0.09, 95% CI: 0.01-0.17, p = 0.032).

**Figure 7.**
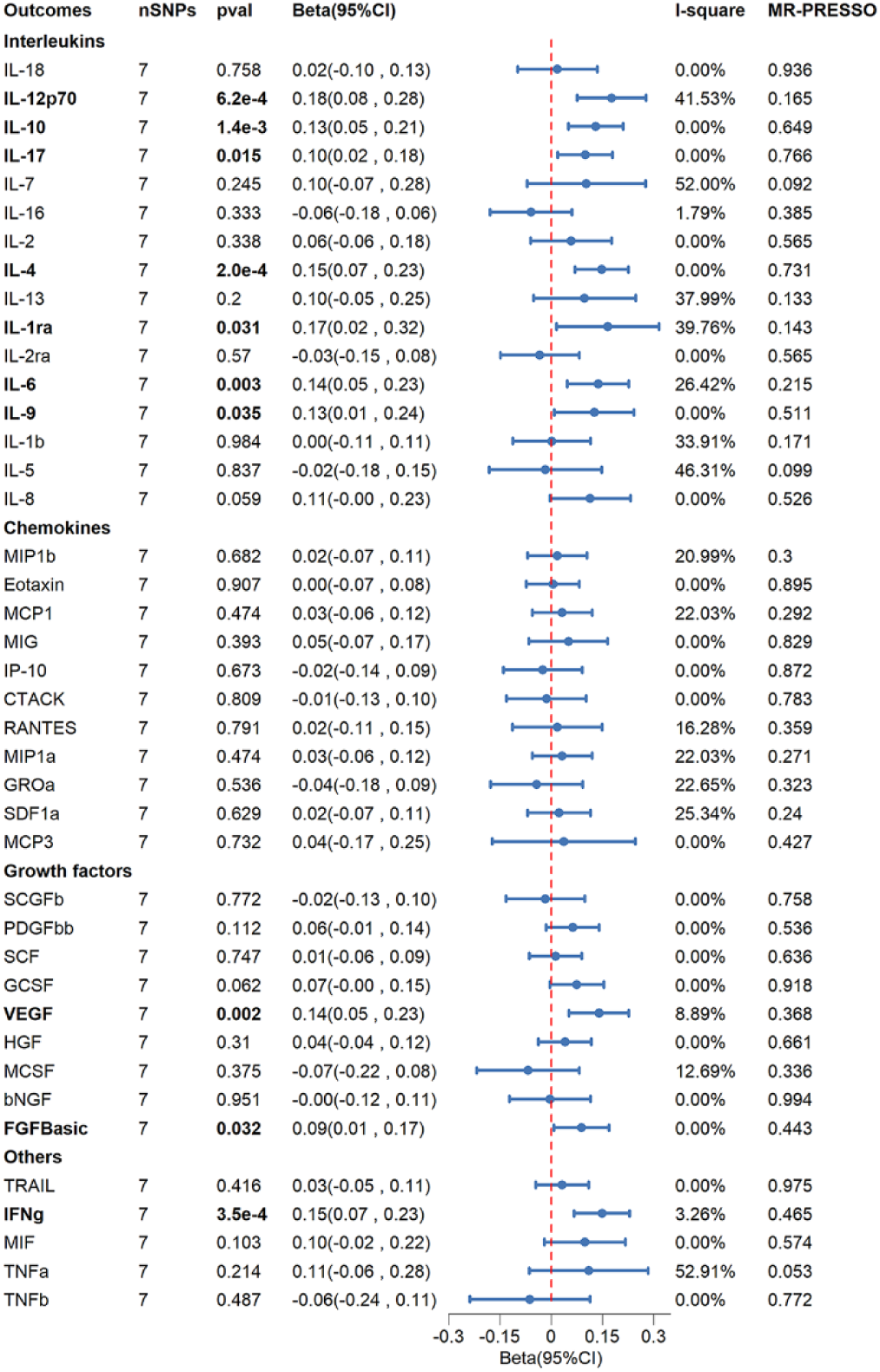
Causalities of intracranial aneurysm (of any type) on 41 inflammatory cytokines. Beta and 95% confidence interval (CI) indicate the alteration in the standard deviation of cytokines per log odds rise in intracranial aneurysm (IA). Significance was determined using a p-value threshold of 0.05 / 41 = 0.0012 following Bonferroni correction. The inverse variance weighted method was applied consistently across all groups.

Using the Weighted Median method, we obtained similar results for IL-12p70 (Beta: 0.15, 95% CI: 0.03-0.26, p = 0.011), IL-4 (Beta: 0.13, 95% CI: 0.03-0.22, p = 0.009), IFNg (Beta: 0.11, 95% CI: 0.01-0.22, p = 0.036), and VEGF (Beta: 0.15, 95% CI: 0.04-0.26, p = 0.009). Moreover, all the results of both the Weighted Median method and the MR-Egger method exhibit a similar estimate trend to the IVW method (Supplementary Table S9). The scatter plots and funnel plots of MR analyses are shown in Figure 8 and Figure 9.

**Figure 8.**
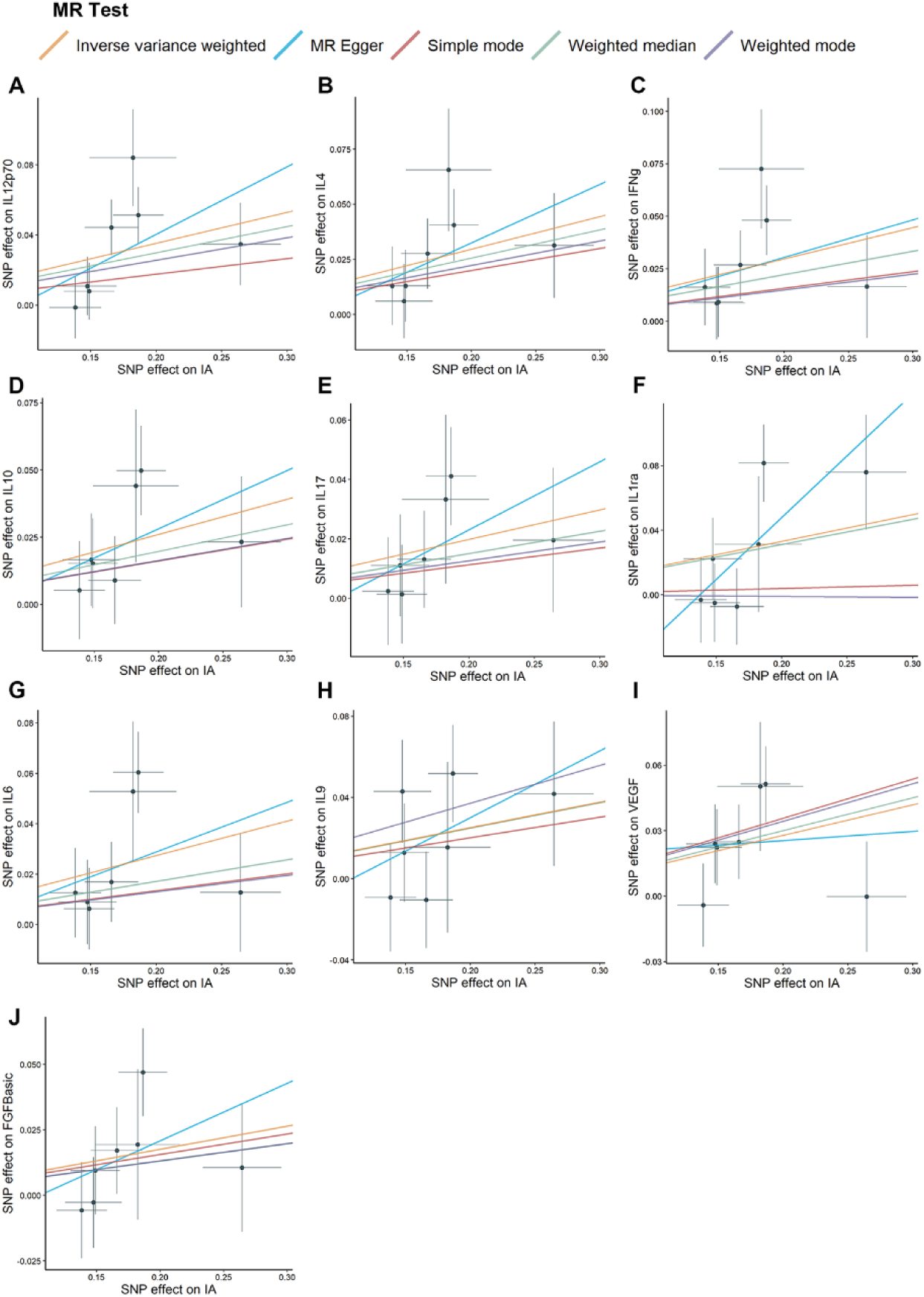
The Mendelian randomization (MR) analyses between intracranial aneurysm (IA) and inflammatory cytokines are depicted through scatter plots. Single nucleotide polymorphisms (SNPs) are depicted as black dots, with standard error bars representing their estimates of IA risk and cytokine levels. The causality is represented by the slope of the line. (A-J): IL-12p70, IL-4, IFNg, IL-10, IL-17, IL-1ra, IL-6, IL-9, VEGF, and FGFBasic.

**Figure 9.**
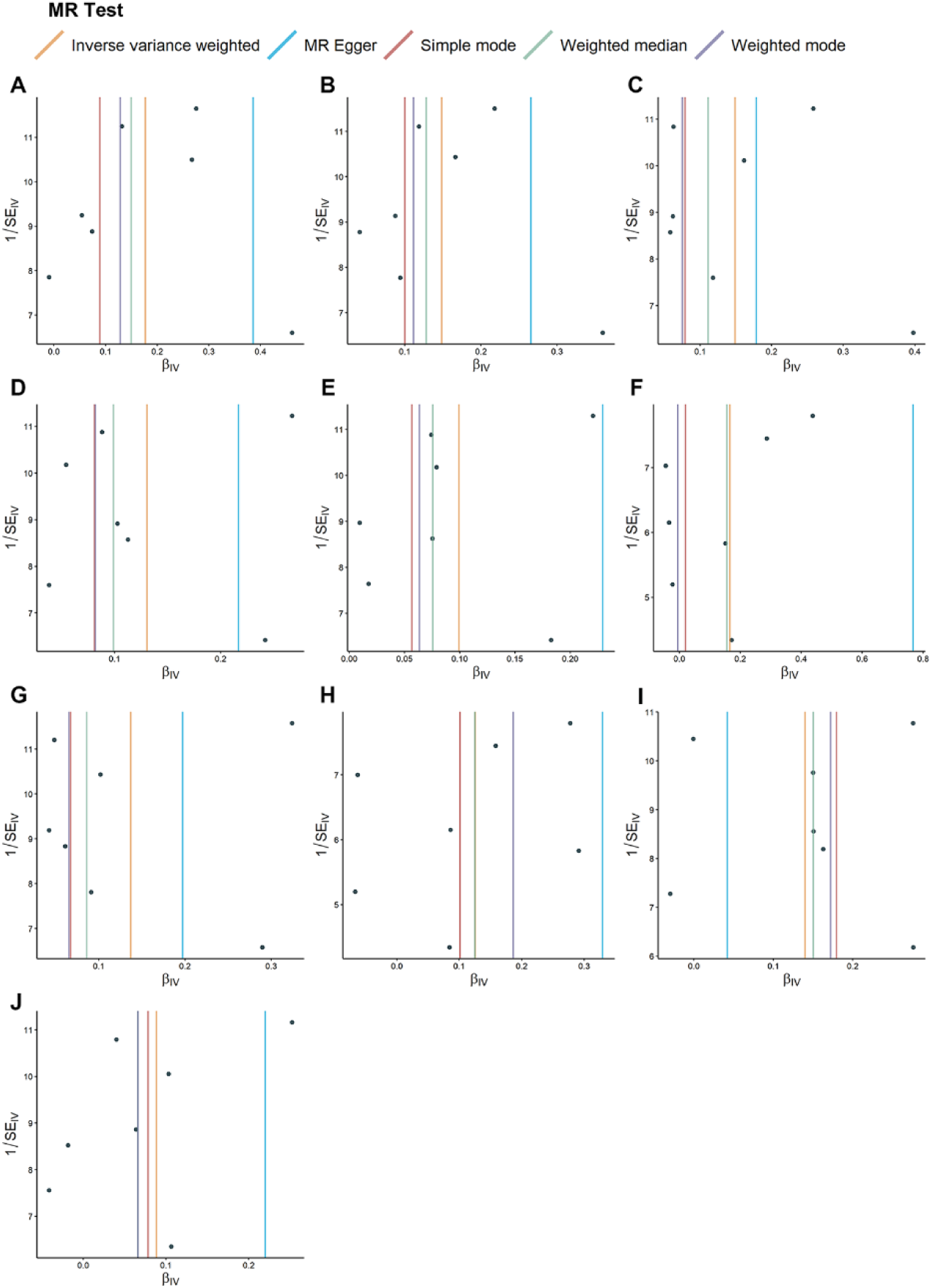
Funnel plots illustrate the Mendelian randomization (MR) analyses between intracranial aneurysm (IA) and specific cytokines. The MR estimation of IA single nucleotide polymorphisms with cytokines versus the reciprocal of the standard error (1/SEIV). (A-J): IL-12p70, IL-4, IFNg, IL-10, IL-17, IL-1ra, IL-6, IL-9, VEGF, and FGFBasic.

## 4. Discussion

This MR study investigated the correlations between inflammatory cytokines (including interleukins, growth factors, chemokines, and others) and UIA or aSAH. The results indicated that cytokines may participate in regulating the formation and development of IA. IP-10 has been identified as a suggestively protective factor in the pathogenesis of UIA. The elevation of IL-10 and VEGF circulating concentrations have a suggestive causality with the increased risk of aSAH. Conversely, FGFBasic and TRAIL exhibit potential protective effects against IA rupture. When IA was considered the exposure, we observed significantly increased circulating concentrations of IL-12p70, IL-4, and IFNg. Simultaneously, there was a suggestively significant increase in IL-10, IL-17, IL-1ra, IL-6, IL-9, VEGF, and FGFBasic concentrations with the onset of IA. The forest plots and leave-one-out sensitivity analyses in this study are presented in supplementary materials (Supplementary Figure S2-S6). Our results indicate that various cytokines have potential regulatory effects on different types of IA, and the invasion of IA can significantly increase the level of circulating cytokines. The MR results suggest a subtle regulatory balance exists between inflammatory cytokines and the pathogenesis and progression of IA.

Several retrospective studies have suggested that the interaction of immune cells, cytokines, and the inflammatory response may exacerbate IA instability and result in a poor prognosis (20-22). The brain injury caused by a ruptured IA is multifaceted and frequently involves immune-inflammatory processes (20,23,24). Therefore, addressing the inflammatory aspect has substantial potential for preventing the progression of IA and mitigating secondary damage resulting from aneurysm rupture. However, most observational studies are either cross-sectional or case-control in nature. Due to the influence of various confounding factors, these studies can only establish associations between cytokines and IA, making it challenging to determine causality (25). Thus, we explored the causal correlation between inflammatory cytokines and IA by utilizing the random distribution of effective genetic instruments based on MR assumptions.

IP-10, also known as CXCL10, is an interferon-γ-induced chemokine that can attract monocytes/macrophages and T cells (26). The results of the MR analysis reveal a suggestively significant negative correlation between the genetically predicted levels of IP-10 and the risk of UIA. An observational study observed an increased level of IP-10 within the cerebral aneurysm cavity compared to the femoral artery, indicating a potential association between IP-10 and the pathogenesis of IA (27). However, the underlying mechanism remains incompletely comprehended. The imbalance of CD4+ T cells is considered one of the factors contributing to the heightened inflammatory state in IA (28). Recent research has uncovered that IP-10 contributes to the repair of human cardiomyocytes by binding CXCR3 to stimulate the transportation of CD4 + Tem cells (29). In our study, increased levels of IP-10 were correlated with a decreased risk of UIA. We hypothesize that the previously observed elevation of IP-10 within IA cysts may be attributed to a negative feedback mechanism by which the vascular wall protects against IA lesions. IP-10 could potentially contribute to the resolution of aneurysm wall inflammation by facilitating the transport of anti-inflammatory macrophages and CD4+ Tem cells, thereby reducing the phenotypic transformation of VSMCs and inhibiting the progression of IA. However, further research is needed to verify this regulatory mechanism.

We observed that FGFBasic and TRIAL can potentially mitigate the risk of aSAH. FGFBasic, also known as FGF2, is a fundamental fibroblast growth factor with mitogenic and angiogenic properties. The deficiency of VSMCs in aneurysms is critical to IA rupture. Previous studies have demonstrated that intravenous injection with FGFBasic in rats retains more VSMCs in the intima of IA walls and enhances aneurysm stability (30). Research on human cerebral aneurysm walls has indicated that the activation of FGFBasic stimulates VSMC proliferation and prevents apoptosis (31). The MR analysis results provide additional confirmation of the beneficial effect of FGFBasic in preventing IA rupture. FGFBasic enhances IA stability and reduces the likelihood of rupture by promoting the proliferation of VSMCs in the IA wall. Tumor Necrosis Factor-Related Apoptosis-Inducing Ligand (TRAIL) is widely expressed in VSMCs. TRIAL induces apoptosis in cancer or transformed cells but not normal cells (32). The limited observational research on TRIAL in IA has led to a debate on its significance in regulating aneurysm stability. TRIAL exhibits immunosuppressive and immunomodulatory functions, promoting early resolution of inflammation and stimulating systemic VSMC proliferation (33-36). Our findings suggest that the upregulation of TRIAL potentially reduces the risk of IA rupture. The protective effect of TRIAL may be associated with the early resolution of inflammation and the stimulation of VSMC proliferation in the aneurysm wall. Further experimental exploration is warranted in the future.

Consistent with most observational studies, MR analysis indicated that VEGF and IL10 were risk factors for aSAH. VEGF is a heparin-binding protein that regulates pathological angiogenesis by inducing vascular endothelial cell proliferation and migration, mediating endothelial injury in the IA wall (37,38). Observational studies have reported elevated VEGF levels in ruptured aneurysm tissue of aSAH patients and cerebrospinal fluid of UIA patients (39,40). Combined with our findings, it appears that VEGF primarily affects the process of aneurysm wall remodeling with IA. VEGF may initially induce damage to endothelial cells in the aneurysm wall, subsequently affecting the structural stability of the inner and middle membranes, finally resulting in the rupture of IA. Aneurysmal wall enhancement (AWE) is associated with inflammation and vulnerability of IA (41). An observational study noted that in unruptured aneurysm cavities, the increase in IL10 was related to the low level of AWE, and aneurysms with low AWE were more unstable (42). Our results reveal that the upregulation of IL10 in circulation leads to an increased risk of aSAH, potentially associated with the low AWE caused by IL10 in vascular remodeling (43). Notably, the results indicate a bidirectional causality between VEGF and IL10 with aSAH, suggesting a possible mechanism of injury including positive feedback. Therefore, targeted therapy for VEGF and IL10 may be crucial in preventing IA rupture.

The process of cerebral aneurysm wall remodeling also involves regulating inflammatory cytokines. Identifying these affected inflammatory factors can enhance our comprehension of the regulatory mechanism of IA inflammation. Previous observational studies have demonstrated that the levels of IL-12p70, IL-4, IFNg, IL-10, IL-17, IL-1ra, IL-6, VEGF, and FGFBasic are positively correlated with the pathogenesis of IA, which is consistent with our findings (30,39,40,44,45). However, there still needs to be an observational study to clarify the association between IL9 and IA. Our results suggest these cytokines have some potential functions in the context of IA. Among them, IL-10, VEGF, and FGFBasic directly impact the prognosis of IA, while IL-12p70, IL-4, IFNg, IL-17, IL-1ra, IL-6, and IL-9 may serve as intermediate factors in regulating IA inflammation.

Based on MR analysis, we have explored causalities between inflammatory cytokines and IA. However, there are still some limitations in this study. First, MR analysis relies on three basic assumptions, but the independence assumption and exclusion restriction cannot be entirely confirmed. Secondly, the conclusions of this study apply to individuals of European ancestry, and caution is advised when extrapolating these results to other racial groups. Lastly, the analysis was restricted to a limited set of cytokines. Further research is warranted to validate and translate our findings into clinical applications for treating and preventing IA.

## 5. Conclusions

In summary, our MR study indicates that IP-10 may function as a protective factor against UIA. FGFBasic and TRAIL demonstrate protective effects in aSAH, while IL-10 and VEGF are identified as the risk factors for IA rupture. Additionally, IL-12p70, IL-4, IFNg, IL-10, IL-17, IL-1ra, IL-6, VEGF, and FGFBasic are upregulated as the consequences of IA and may participate in regulating inflammation in IA pathogenesis.

## Data Availability Statement

The original contributions presented in the study are included in the article/Supplementary Material, further inquiries can be directed to the corresponding author.

## Author Contributions

QZ completed the data analysis and the writing of the paper. TF and TL critically revised the manuscript. LZ, YZ, and XS checked for omissions in the study and provided comments. TW, XK, HL, JL, and YL participated in the formulation of the draft study design. The final manuscript has been read and approved by all authors.

