## Supplementary Figures for "Circulating Inflammatory Cytokines and Risk of Intracranial Aneurysm: A Mendelian-randomization Study"

**Supplementary Figure S1.** Causalities of aSAH on cytokines. Beta and 95% CI indicate the alteration in the standard deviation of cytokines per log odds rise in aSAH. Significance was determined using a p-value threshold of 0.05 / 41 = 0.0012 following Bonferroni correction. The inverse variance weighted method was applied consistently across all groups except for IL-13, IL-1ra, and TNFa, which applied the IVW (multiplicative random effects) method.

| Outcomes | nSNPs | pval | Beta(95%CI) | I-square | MR-PRESSO |
|---|---|---|---|---|---|
| **Interleukins** | | | | | |
| IL-18 | 5 | 0.593 | 0.03(-0.09 , 0.15) | 0.00% | 0.893 |
| **IL-12p70** | 5 | **0.005** | 0.15(0.05 , 0.26) | 43.20% | 0.212 |
| **IL-10** | 5 | **0.01** | 0.11(0.03 , 0.20) | 0.66% | 0.43 |
| IL-17 | 5 | 0.053 | 0.08(-0.00 , 0.17) | 1.55% | 0.446 |
| IL-7 | 5 | 0.555 | 0.05(-0.12 , 0.23) | 48.03% | 0.18 |
| IL-16 | 5 | 0.62 | -0.04(-0.18 , 0.10) | 21.30% | 0.32 |
| IL-2 | 5 | 0.723 | 0.02(-0.11 , 0.16) | 17.69% | 0.297 |
| **IL-4** | 5 | **0.002** | 0.13(0.05 , 0.21) | 0.00% | 0.692 |
| IL-13 | 5 | 0.302 | 0.10(-0.09 , 0.29) | 58.39% | 0.099 |
| IL-1ra | 5 | 0.17 | 0.14(-0.06 , 0.34) | 62.26% | 0.058 |
| IL-2ra | 5 | 0.634 | -0.03(-0.15 , 0.09) | 0.00% | 0.751 |
| IL-6 | 5 | 0.067 | 0.11(-0.01 , 0.23) | 54.73% | 0.103 |
| IL-9 | 5 | 0.193 | 0.08(-0.04 , 0.21) | 7.87% | 0.383 |
| IL-1b | 5 | 0.621 | -0.03(-0.16 , 0.10) | 45.67% | 0.164 |
| IL-5 | 5 | 0.643 | -0.04(-0.23 , 0.14) | 54.64% | 0.094 |
| IL-8 | 5 | 0.255 | 0.08(-0.06 , 0.21) | 13.06% | 0.362 |
| **Chemokines** | | | | | |
| MIP1b | 5 | 0.81 | 0.01(-0.10 , 0.13) | 49.80% | 0.152 |
| Eotaxin | 5 | 0.767 | 0.01(-0.07 , 0.09) | 0.00% | 0.95 |
| MCP1 | 5 | 0.144 | 0.06(-0.02 , 0.14) | 0.00% | 0.799 |
| MIG | 5 | 0.562 | 0.04(-0.08 , 0.16) | 0.00% | 0.88 |
| IP-10 | 5 | 0.792 | -0.02(-0.14 , 0.10) | 0.00% | 0.826 |
| CTACK | 5 | 0.59 | -0.03(-0.16 , 0.09) | 0.00% | 0.711 |
| RANTES | 5 | 0.726 | -0.03(-0.17 , 0.12) | 25.16% | 0.319 |
| MIP1a | 5 | 0.257 | 0.09(-0.07 , 0.25) | 37.14% | 0.201 |
| GROa | 5 | 0.956 | -0.00(-0.13 , 0.12) | 0.00% | 0.528 |
| SDF1a | 5 | 0.492 | 0.03(-0.06 , 0.12) | 10.12% | 0.376 |
| MCP3 | 5 | 0.938 | -0.01(-0.26 , 0.24) | 26.06% | 0.264 |
| **Growth factors** | | | | | |
| SCGFb | 5 | 0.564 | -0.04(-0.16 , 0.09) | 0.00% | 0.551 |
| PDGFbb | 5 | 0.389 | 0.04(-0.05 , 0.13) | 20.15% | 0.332 |
| SCF | 5 | 0.823 | -0.01(-0.09 , 0.07) | 0.00% | 0.531 |
| GCSF | 5 | 0.168 | 0.06(-0.02 , 0.14) | 0.00% | 0.662 |
| **VEGF** | 5 | **0.039** | 0.11(0.01 , 0.22) | 32.53% | 0.27 |
| HGF | 5 | 0.252 | 0.05(-0.03 , 0.13) | 0.00% | 0.708 |
| MCSF | 5 | 0.296 | -0.10(-0.28 , 0.09) | 37.39% | 0.235 |
| bNGF | 5 | 0.865 | -0.01(-0.13 , 0.11) | 0.00% | 0.966 |
| FGFBasic | 5 | 0.074 | 0.09(-0.01 , 0.18) | 22.72% | 0.316 |
| **Others** | | | | | |
| TRAIL | 5 | 0.4 | 0.03(-0.05 , 0.12) | 0.00% | 0.967 |
| **IFNg** | 5 | **0.004** | 0.12(0.04 , 0.21) | 1.72% | 0.442 |
| MIF | 5 | 0.413 | 0.05(-0.07 , 0.18) | 0.00% | 0.538 |
| TNFa | 5 | 0.64 | 0.05(-0.15 , 0.25) | 61.28% | 0.079 |
| TNFb | 5 | 0.683 | -0.04(-0.22 , 0.15) | 0.00% | 0.76 |

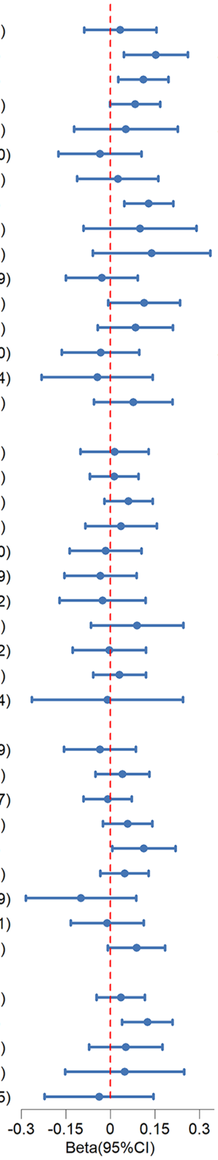

**Supplementary Figure S2.** Forest plot and leave-one-out sensitivity analyses of MR analysis between inflammatory cytokines and UIA.

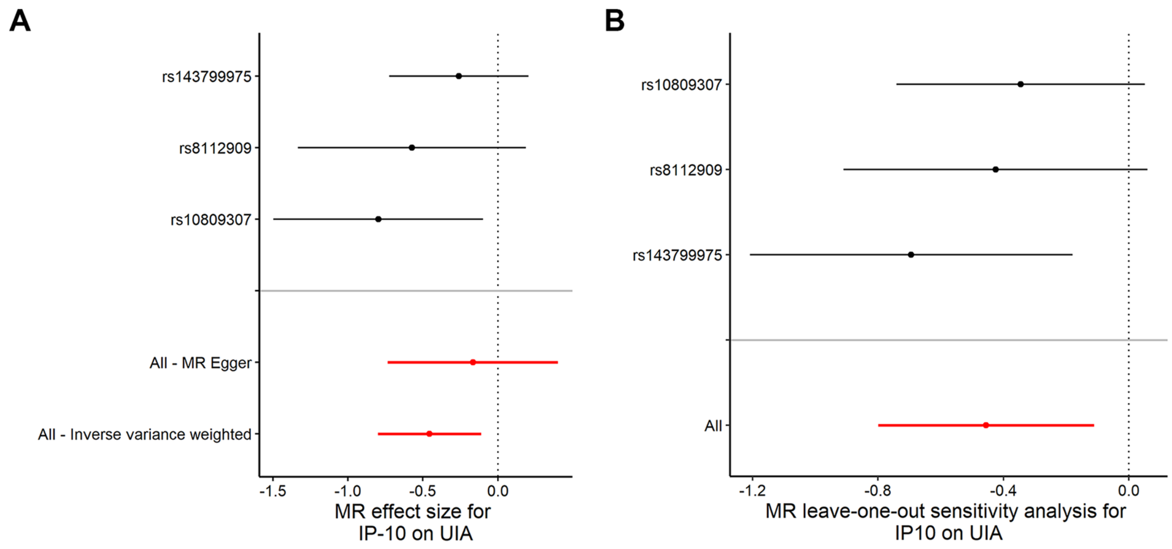

**Supplementary Figure S3.** Forest plots of MR analysis between inflammatory cytokines and aSAH. (A-D): IL-10, VEGF, FGFBasic, and TRAIL.

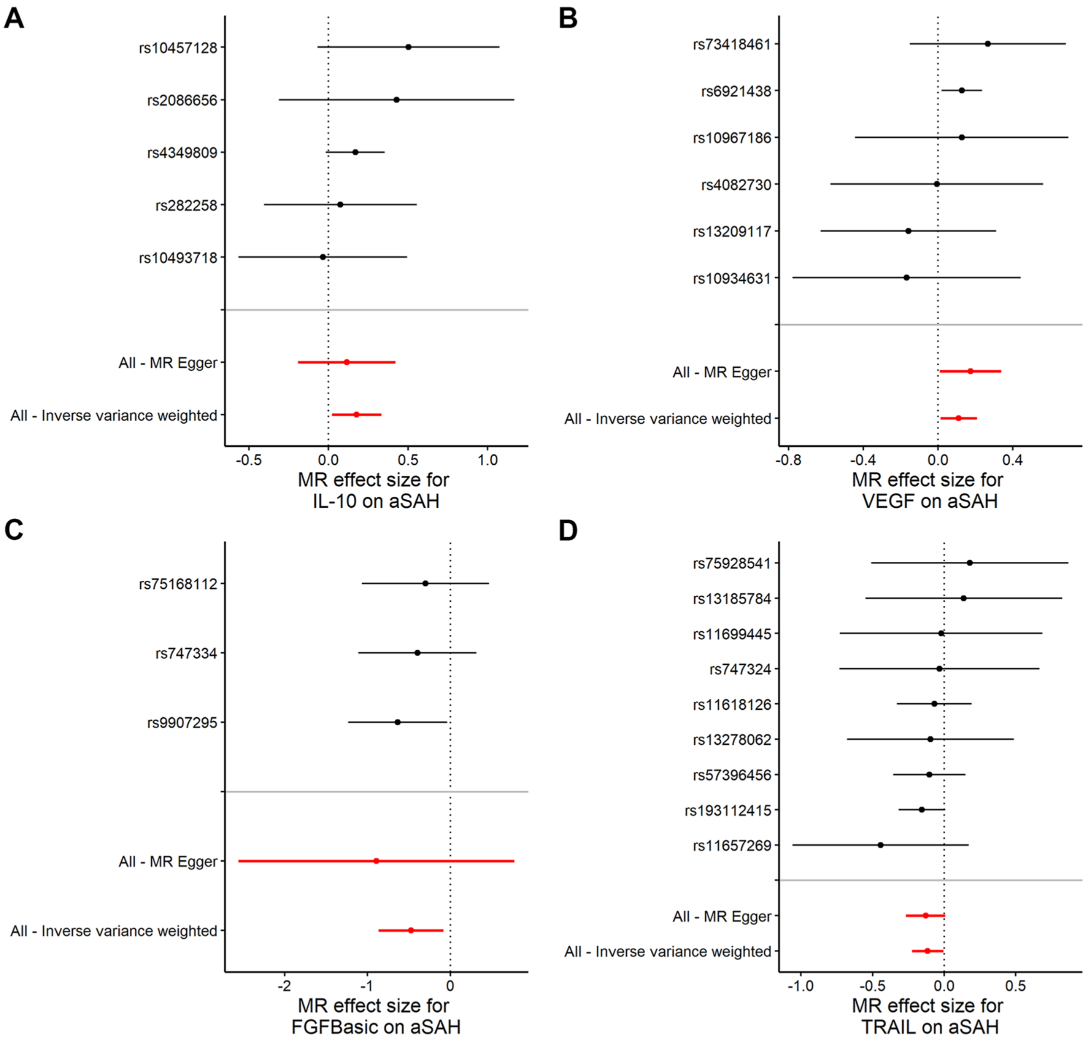

**Supplementary Figure S4.** Leave-one-out sensitivity analyses of MR analysis between inflammatory cytokines and aSAH. (A-D): IL-10, VEGF, FGFBasic, and TRAIL.

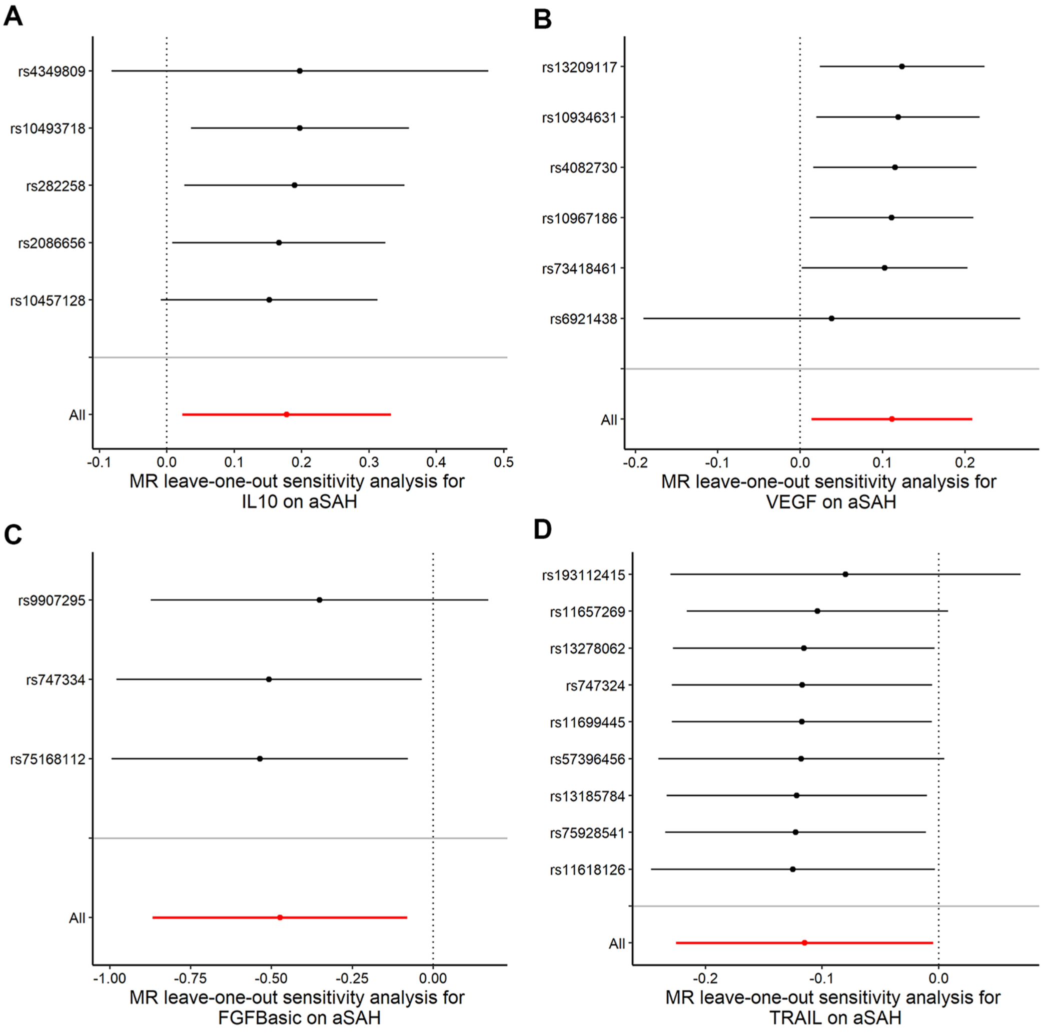

**Supplementary Figure S5.** Forest plots of MR analysis between IA and inflammatory cytokines. (A-J): IL-12p70, IL-4, IFNg, IL-10, IL-17, IL-1ra, IL-6, IL-9, VEGF, and FGFBasic.

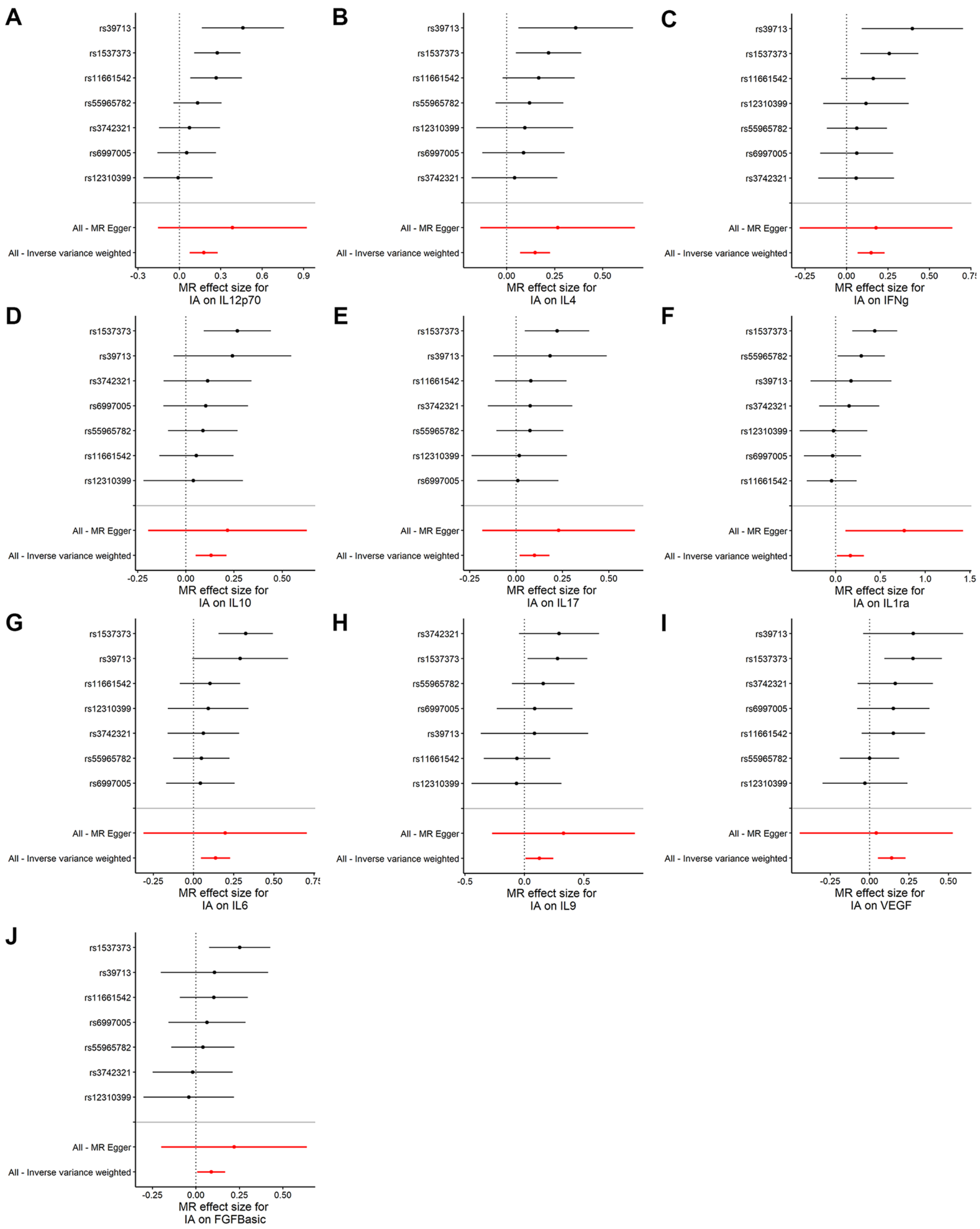

**Supplementary Figure S6.** Leave-one-out sensitivity analyses of MR analyses between IA and inflammatory cytokines. (A-J): IL-12p70, IL-4, IFNg, IL-10, IL-17, IL-1ra, IL-6, IL-9, VEGF, and FGFBasic.

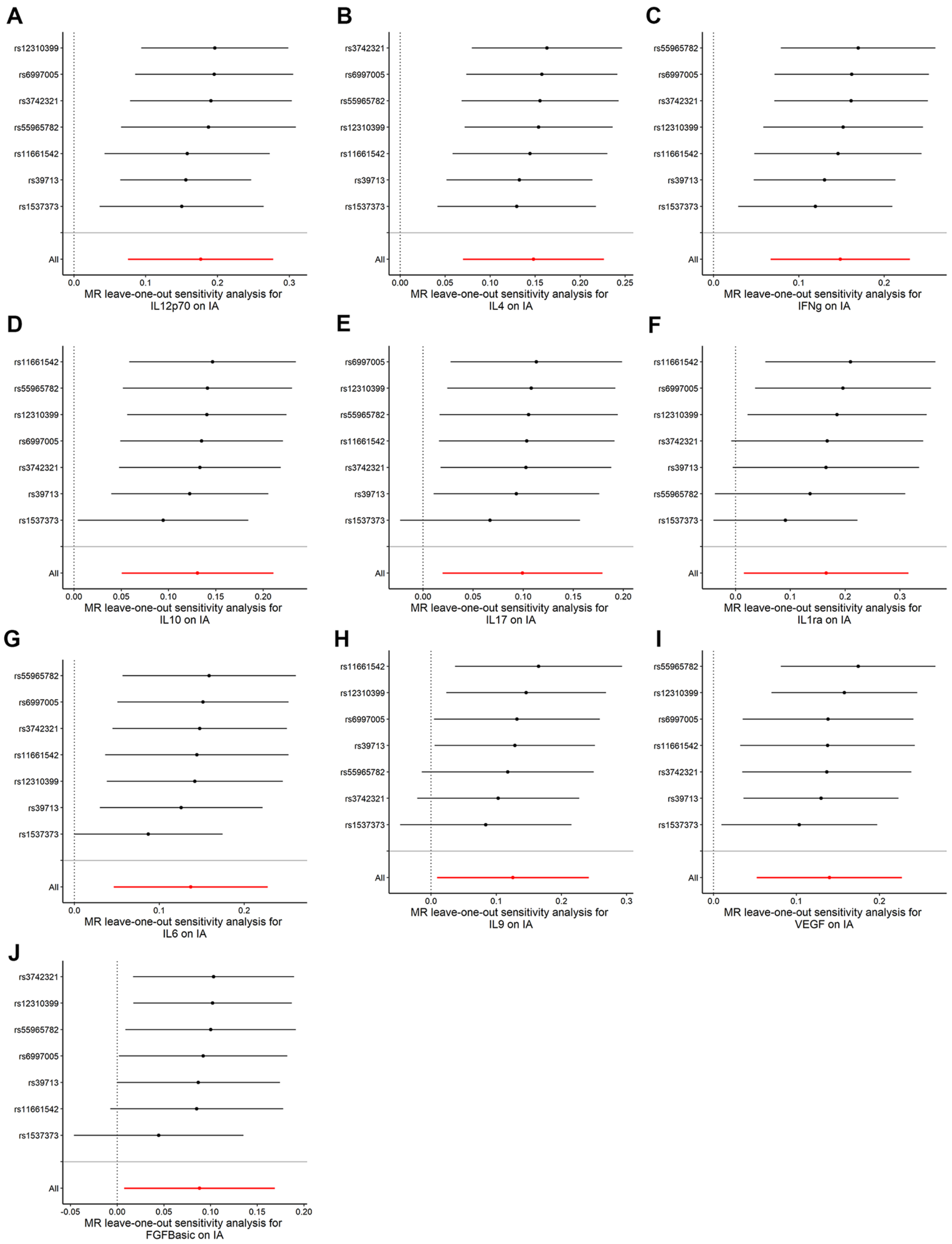
